# Association between maternal organophosphate flame retardants (OPFRs) metabolite levels during pregnancy and internalizing and externalizing behaviors in children at 2-5 years

**DOI:** 10.1101/2025.10.28.25338905

**Authors:** Yaoyilian Cheng, Sarina Abrishamcar, Tszshan Ma, Stephanie M. Eick, Dana Boyd Barr, Nadia Hoffman, Syam S Andra, Robert O Wright, Heather J Zar, Dan J Stein, Anke Huels

**Affiliations:** Department of Epidemiology, Rollins School of Public Health, Emory University, Atlanta, GA, USA; Gangarosa Department of Environmental Health, Rollins School of Public Health, Emory University, Atlanta, GA, USA; Neuroscience Institute, University of Cape Town, Cape Town, South Africa; Department of Psychiatry and Mental Health, University of Cape Town, Cape Town, South Africa; Department of Environmental Medicine, Icahn School of Medicine at Mount Sinai, New York, NY 10029, USA; Department of Pediatrics and Child Health, Red Cross War Memorial Children’s Hospital, University of Cape Town, Cape Town, South Africa; South African Medical Research Council (SAMRC) Unit on Child and Adolescent Health, University of Cape Town, Cape Town, South Africa; South African Medical Research Council (SAMRC) Unit on Risk and Resilience in Mental Disorders, University of Cape Town, Cape Town, South Africa; Department of Biostatistics and Bioinformatics, Rollins School of Public Health, Emory University, Atlanta, GA, USA

## Abstract

**Background:** Organophosphorus flame retardants (OPFRs) metabolites are frequently detected in human samples. Their structural similarity to polybrominated diphenyl ethers raises concerns about potential neurotoxicity. Animal and limited human studies suggest prenatal exposure to OPFRs may affect offsprings’ neurobehavioral problems.

**Objectives:** We investigated the association between maternal OPFRs exposure and neurobehavioral problems in children aged two to five years from the South African Drakenstein Child Health Study (N=705).

**Methods:** We measured urinary metabolites of OPFRs, bis(1,3-dichloro-2-propyl) phosphate (BDCPP) and diphenyl phosphate (DPHP) during the 2^nd^ trimester of pregnancy or at birth, using a targeted multiclass assay. Childhood behaviors were repeatedly assessed at 24, 42 and 60 months using the Child Behavior Checklist (CBCL). After natural log transformation of metabolite concentrations, we used linear regression to analyze the association between BDCPP and DPHP levels and internalizing/externalizing behaviors t-scores, adjusting for maternal age, body mass index, ancestry, and socioeconomic status.

**Results:** At 24 months, each standard deviation (SD) increase in log-BDCPP levels was associated with higher externalizing (1.26, 95% CI: 0.30, 2.21) and internalizing (1.14, 95% CI: 0.02, 2.25) subscores. Similarly, one SD increase in log-DPHP was associated with higher externalizing (+1.19, 95% CI: 0.23, 2.16) and internalizing (1.00, 95% CI: −0.12, 2.13) subscores. Only DPHP levels were associated with internalizing subscores at 60 months.

**Conclusion:** We observed a positive association between prenatal metabolite levels of OPFRs and neurobehavioral problems among children at age two, raising concerns about prenatal chemical exposures and awareness of developing interventions to promote healthier developmental outcomes for children.

## Introduction

Organophosphate flame retardants (OPFRs) are a group of emerging compounds increasingly used as alternatives to traditional flame retardants, such as polybrominated diphenyl ethers (PBDEs).

The environmental persistence and ecotoxicity of PBDEs have led to their phase-out and replacement with OPFRs. ^1^ Since the early 2000s, following the ban on PBDEs, the production and consumption of OPFRs have surged. OPFRs are widely used in products such as furniture, textiles, plastics, electronics, and other materials to reduce flammability. ^2–4^ Additionally, OPFRs are used in some nail polishes. ^5^ As a result, OPFRs are frequently detected in indoor air, dust, and on household surfaces, making inhalation and dermal contact common exposure routes. Furthermore, dietary intake, particularly from food packaging and processed goods, can serve as another pathway for human exposure. ^6–8^ Metabolites of OPFRs—particularly bis(1,3-dichloro-2-propyl) phosphate (BDCPP) and diphenyl phosphate (DPHP)—have been consistently detected in a variety of human samples, particularly in urine. ^9^ Long-term assessments in the United States have indicated an upward temporal trend, with concentrations of BDCPP and DPHP in urine samples significantly increasing since 2002, particularly in the Northeastern United States. ^10^

The growing presence and ubiquity of OPFRs in the environment raises serious concerns about their potential health impacts. Toxicological studies have demonstrated that, similar to organophosphate pesticides and PBDEs, OPFRs have the ability to disrupt the endocrine system in humans and experimental animals. ^2^ This disruption can lead to harmful reproductive and neurodevelopmental effects, suggesting that exposure to OPFRs may have transgenerational consequences. ^11,12^ One of the most significant concerns related to OPFR exposure during pregnancy is the potential effect on neurodevelopment. A review of thirteen U.S. epidemiologic studies showed that levels of BDCPP and DPHP tended to be higher among pregnant women compared with other populations raising concerns about increased exposure to OPFRs during pregnancy. ^10^ This is particularly alarming because, according to the Developmental Origins of Health and Disease (DoHaD) hypothesis, certain chemicals to which pregnant women are exposed can cross the placenta and affect fetal development, leading to adverse pregnancy outcomes and developmental problems later in life. ^13^ Several studies have shown that increased OPFR metabolite levels during pregnancy are associated with adverse neurodevelopment outcomes, including lower intelligence quotient, poor working memory, delayed psychomotor development, and impaired early language abilities in children. ^14–16^ These findings raise concern about the potential of OPFRs to contribute to neurodevelopmental delays and cognitive deficits, which could have long-term consequences on a child’s learning and social adaptation.

Despite the growing body of evidence of the negative impact of prenatal OPFR exposure on neurobehavioral development in offspring, most studies to-date are cross-sectional and based in high-income countries, ^17,18^ limiting their generalizability. A significant gap remains regarding the impact of prenatal OPFRs exposure in low- and middle-income countries (LMICs), where exposure sources and vulnerabilities may differ. In South Africa, for example, OPFRs are prevalent in both indoor and outdoor environments due to widespread use in consumer products, posing an exposure risk to pregnant individuals. ^19,20^ Thus, it is necessary to conduct research on the effects of prenatal OPFRs exposure on neurobehavior in this context.

To address this gap, we used data from 705 mothers-child pairs enrolled in the South African Drakenstein Child Health Study to investigate the association between prenatal OPFR metabolite levels and neurobehavioral problems in early childhood. Our research aimed to assess the association of OPFR metabolite levels with internalizing and externalizing behaviors by analyzing maternal urine samples collected either during 2^nd^ trimester pregnancy or at birth, along with behavioral assessments of children measured at 24, 42, and 60 months repeatedly. We hoped to provide a better understanding of how prenatal OPFR exposures contribute to risks of neurobehavioral developmental problems in offspring with unique environmental conditions and how the effect changes as children grow.

## Methods

### Study design and population

The Drakenstein Child Health Study (DCHS) is a prospective population based birth cohort located in a peri-urban area of Drakenstein, Cape Town, South Africa, aiming to investigate the risk factors and impact of early exposures on child health. ^21^ Pregnant women were enrolled between 20- and 28-weeks gestation at the Mbekweni or TC Newman clinics from March 2012 to March 2015 and were followed through birth and through childhood. Eligibility criteria included mothers being at least 18 years of age, attending prenatal care at one of the two public primary health clinics, and intending to remain in the area for at least one year. ^21^ Mother-child dyads were followed at multiple postnatal timepoints: 6-10 weeks, 14 weeks, 6 months, 9 months, 12 months, and 6 monthly to annually thereafter. All participants provided written informed consent at enrollment and renewed it annually A total of 1,137 mothers and 1,143 children were originally enrolled in DCHS ^21^. In the present study, we included 705 mother-child pairs with prenatal OPFRs urinary metabolite measurements either during pregnancy or at birth, at least one complete child behavior checklist (CBCL) assessment at 24 months (N =459), 42 months (N=662) or 60 months (N=678), and information on all relevant covariates. The DCHS was approved by the faculty of Health Sciences, Human Research Ethics Committee, University of Cape Town (401/2009), Stellenbosch University (N12/02/0002) and the Western Cape Provincial Health Research committee (2011RP45).

### Prenatal OPFRs exposure assessment

Urinary concentrations of two OPFR metabolites, BDCPP and DPHP were measured in 705 maternal urine samples collected during the second trimester of pregnancy or at birth. Most urine samples (n = 643) were collected either during pregnancy at 2^nd^ trimester (20- and 28-week gestation) while a urine sample was taken at birth for those who were missing a pregnancy sample (n = 62). Urine samples were collected using a clean-catch, spot method and placed into sterile polypropylene collection tubes. The samples were then transported, processed, aliquoted, and stored frozen at the DCHS research laboratory. They were cataloged in the DCHS biorepository using the Freezerworks inventory system. Urinary metabolite measurements were performed by the Human Health Exposure Analysis Resource (HHEAR) Targeted Analysis Laboratory at the Icahn School of Medicine at Mount Sinai in New York, NY, using a previously developed and validated multiclass assay which has been confirmed through proficiency testing and has consistently met acceptance criteria in the bi-annual participation in the German External Quality Assessment Scheme (G-EQUAS; http://www.g-equas.de/) and the Centre de toxicologie du Québec (CTQ) External Quality Assessment Scheme for Organic Substances in Urine (CTQ-OSEQAS; https://www.inspq.qc.ca/en/ctq/eqas/oqesas/description) programs. ^22^ This method employed solid-phase extraction and isotope-dilution liquid chromatography-tandem mass spectrometry. Quality control (QC) includes the use of experimental blanks (both reagent- and matrix-based), matrix spikes at different validation levels (low, medium, and high), investigator-initiated paired duplicate samples that were blinded to the laboratory, HHEAR QC urine pools A and B (three pools for every 100 samples), NIST standard reference materials (SRM 3672 and SRM 3673), and archived proficiency-testing materials, as specified by HHEAR. ^23^ Concentrations of BDCPP and DPHP were log-transformed and scaled for statistical analysis, including both well- and not well-detected metabolites. Concentrations below the limits of detection (LODs) were imputed with the LOD⁄√2 in the analysis. ^24^

### Outcome assessment

Childhood behavior was assessed at 24, 42 and 60 months of age using the pre-school version of the Child Behavior Checklist (CBCL), an instrument widely used and validated to detect behavioral and emotional problems in children and adolescents. ^25^ The CBCL was completed by parents or caregivers. It consists of 113 questions scored on a three-point Likert scale (0=absent, 1= occurs sometimes, 2=occurs often), which can be grouped into internalizing and externalizing behaviors subscores. The internalizing subscores were calculated by the combination of anxious/depressed, withdrawn/depressed, and somatic complaints syndromic scales and the externalizing subscores were calculated based on the rule-breaking and aggressive behavior syndromic scales. ^25^ A higher CBCL score indicates greater severity of behavioral problems. Standardized t-scores for internalizing and externalizing behaviors were used for the statistical analysis.

### Statistical Analysis

We used both unadjusted and adjusted linear regression models to analyze the association between prenatal OPFRs metabolite concentrations and internalizing/externalizing behaviors subscores at 24, 42 and 60 months of age. We used a directed acyclic graph (DAG) to select potential confounders for the linear regression model (Supplemental Figure 1). Covariates adjusted were maternal body mass index (BMI) and maternal age at enrollment, child’s ancestry (Black African or Mixed ancestry), and socioeconomic status (SES). We used child’s ancestry to account for cultural differences that could affect exposure levels. We calculated maternal BMI by heights and weights collected at enrollment. Maternal age and child’s ancestry were self-reported at enrollment. SES was measured by the sum scores of standardized SES components (assets, education, employment and income). Higher sum score indicated higher SES. In our extended models, we further adjusted for creatinine concentration to account for variation in urinary dilution in spot samples.

### Sensitivity analyses

We compared the effect of each chemical on internalizing and externalizing behaviors among the full study sample, which included OPFR metabolites measured during pregnancy as well as at birth, to the subset that included only individuals with biomarker measurements taken during the second trimester of pregnancy. Due to the relatively low detection frequency of BDCPP and DPHP, we also categorized them as binary variables (detected/not detected) to investigate whether there was an association between OPFRs urinary metabolites detection and internalizing and externalizing behaviors. Additionally, we conducted an effect modification analysis for infant sex to evaluate whether associations between OPFRs exposure and childhood behavior differ between males and females.

All the statistical analyses were performed using R version 4.4.1.

## Results

### Study Population

The final study sample consisted of 705 mother-child pairs, of which 459 children had CBCL measurements available at 24 months, 662 at 42 months and 678 at 60 months (Table 1). The mean maternal age at enrollment was 28.0 (SD = 6.0) years, and 46% of the offsprings were female (Table 1). The mean maternal BMI in the second trimester was 26.9 (SD = 6.0) kg/m^2^. Almost half of participants self-reported as Black African (n = 346; 49%) and the remaining participants self-reported as mixed ancestry (n = 359; 51%). Most participants’ highest educational achievement was secondary school (> 6-year education). The lower-bound poverty line for South Africa from 2012 to 2015 was approximately ZAR 541 per person per month to ZAR 613 per person per month. ^26^ South African households had an average annual income of R138,168 in 2015, which breaks down to about R11,514 per month. ^27^ 36% of the participants’ (n = 245) current household income was lower than R1,000 per month. Half of the individuals were unemployed (n = 361, 51%).

**Table 1.** Maternal and children’s characteristics, OPFRs concentrations, and t-score of CBCL at 24, 42 and 60 months in Drakenstein Child Health Study from 2012 to 2015, Cape Town, South Africa.

| Characteristic | Overall<sup>a</sup><br>(N=705) | 24<br>months<br>(N=459) | 42<br>months<br>(N=662) | 60 months<br>(N=678) |
| --- | --- | --- | --- | --- |
| Maternal BMI, mean ± SD | 28.0 ± 6.0 | 28.4 ± 6.2 | 28.2 ± 6.3 | 28.0 ± 6.0 |
| Maternal age, mean ± SD | 26.9 ± 6.0 | 27.0 ± 5.9 | 26.8 ± 5.9 | 26.9 ± 6.0 |
| Ancestry, *n* (%) |  |  |  |  |
| Black African | 346<br>(49%) | 223<br>(49%) | 329<br>(50%) | 332 (49%) |
| Mixed Ancestry | 359<br>(51%) | 236<br>(51%) | 333<br>(50%) | 346 (51%) |
| Highest maternal educational achievement, *n* (%) |  |  |  |  |
| Primary | 60 (9%) | 41 (8.9%) | 57 (8.6%) | 55 (8.1%) |
| Some Secondary | 371<br>(53%) | 236<br>(51%) | 354<br>(53%) | 359 (53%) |
| Completed Secondary | 236<br>(33%) | 153<br>(33%) | 215<br>(32%) | 229 (34%) |
| Any Tertiary | 38 (5%) | 29 (6.3%) | 36 (5.4%) | 35 (5.2%) |
| Current household income, *n* (%) |  |  |  |  |
| <R1,000/m | 255<br>(36%) | 161<br>(35%) | 244<br>(37%) | 245 (36%) |
| R1,000-5,000/m | 357<br>(51%) | 235<br>(51%) | 332<br>(50%) | 345 (51%) |
| >R5,000/m | 93 (13%) | 63 (14%) | 86 (13%) | 88 (13%) |
| Current parental employment, *n* (%) |  |  |  |  |
| Not Working | 361<br>(51%) | 229<br>(50%) | 432<br>(52%) | 348 (51%) |
| Working | 344<br>(49%) | 230<br>(50%) | 320<br>(48%) | 330 (49%) |
| Number of assets in household, mean ± SD | 7 ± 2 | 7 ± 2 | 7 ± 2 | 7 ± 2 |
| SES sum score, mean ± SD | 0.2 ± 2.1 | 0.26 ± 2.16 | 0.14 ± 2.11 | 0.19 ± 2.11 |
| Child sex, *n* (%) |  |  |  |  |
| Female | 325<br>(46%) | 203<br>(44%) | 305<br>(46%) | 313 (46%) |
| Male | 380<br>(54%) | 256<br>(56%) | 357<br>(54%) | 365 (54%) |
| BDCPP concentration (ng/ml),<br>median ± IQR | 0.33 ±<br>1.12 | 0.28 ±<br>0.82 | 0.31 ±<br>1.02 | 0.32 ± 1.07 |
| DPHP concentration (ng/ml), median<br>± IQR | 1.15 ±<br>1.79 | 1.00 ±<br>1.80 | 1.11 ±<br>1.65 | 1.12 ± 1.76 |
| Creatinine concentration<sup>b</sup> (mg/dL),<br>median ± IQR | 99 ± 95 | 99 ± 87 | 97 ± 94 | 99 ± 97 |
| CBCL scores |  |  |  |  |
| *t*-score of Externalizing<br>behaviors, mean ± SD | - | 46 ± 11 | 42 ± 11 | 42 ± 11 |
| *t*-score of Internalizing<br>behaviors, mean ± SD | - | 46 ± 12 | 41 ± 10 | 45 ± 13 |

<sup>a</sup> Analysis sample with CBCL measurements from at least one of the three timepoints
<sup>b</sup> Creatinine concentrations are missing for 3 of the 705 participants.
Abbreviations: BMI: body mass index; SES: socioeconomic status; BDCPP: bis(1,3-dichloro-2-propyl) phosphate; DPHP: diphenyl phosphate; CBCL: Child Behavior Checklist.

The mean t-score of externalizing behaviors among all children at 24 months was 46 (SD = 11) and it decreased slightly to 42 (SD = 11) when c^27^hildren turned 42 and 60 months. The mean t-score of internalizing behaviors was also 46 (SD = 12) at 24 months while it changed to 41 (SD =10) at 42 months and 45 (SD = 13) at 60 months (Table 1). The median concentration of BDCPP was 0.33 (IQR = 1.12) ng/ml and the median concentration of DPHP was 1.15 (IQR = 1.70) ng/ml in the study sample. These concentrations showed little variation among subsamples with measurements of CBCL at 24, 42, and 60 months only (Table 1). In the urine samples used in our analysis (N =705), 41% and 76% of the samples had detectable (>0.5 ng/mL) levels of BDCPP and DPHP, respectively.

### Association Between OPFR Metabolite Levels and CBCL Subscores

In adjusted linear regression models for outcomes measured at 24 months (n = 459), each standard deviation (SD) increase in the natural log-transformed BDCPP metabolite concentrations were associated with significantly higher t-scores for internalizing and externalizing behaviors after adjusting for maternal BMI, maternal age, ancestry, and SES (Supplemental Table 2, Figure 1).

**Figure 1.**
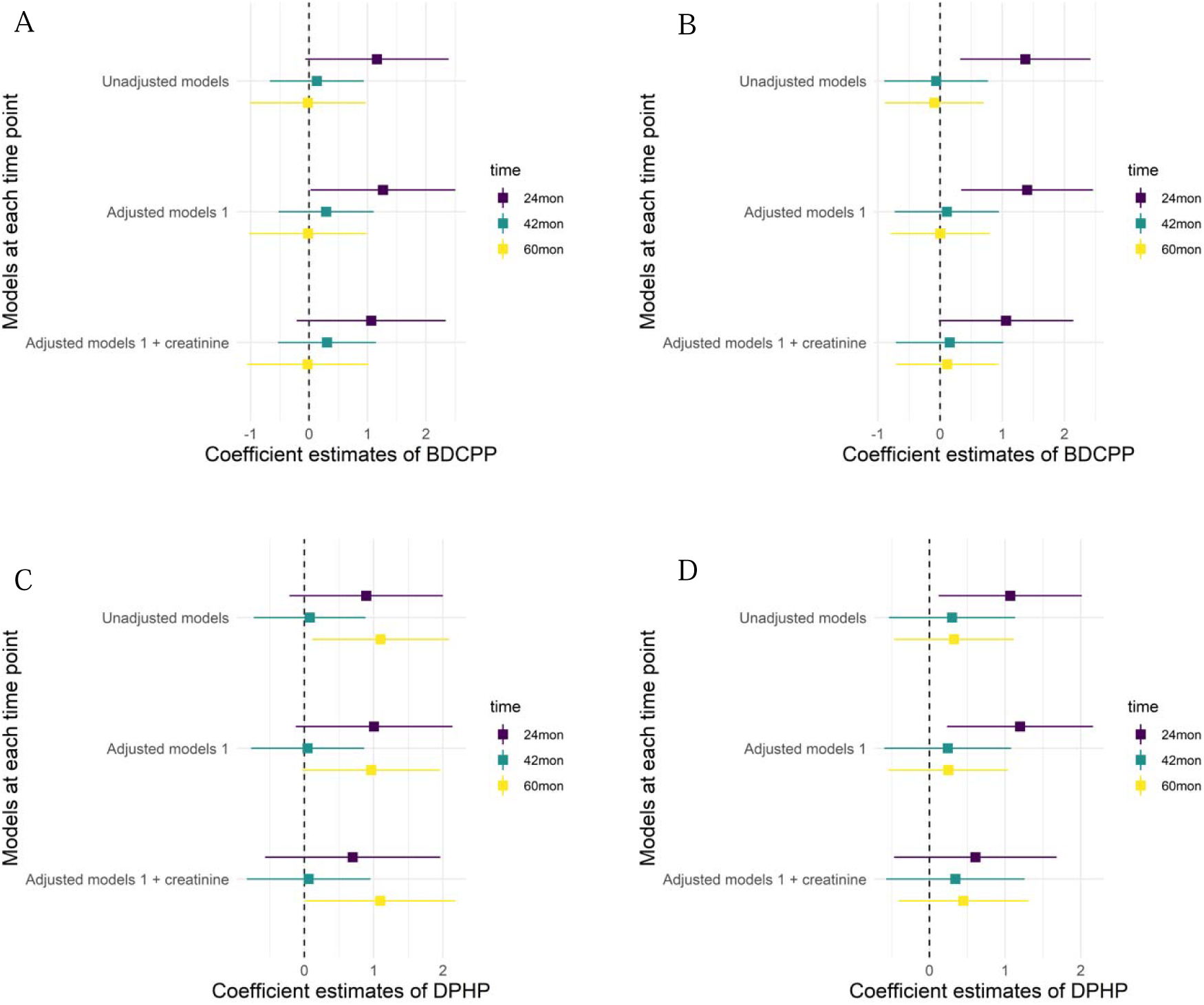
Association between prenatal OPFR metabolite levels and internalizing and externalizing behaviors at 24, 42, and 60 months in Drakenstein Child Health Study from 2012 to 2015, Cape Town, South Africa. (A) Association between BDCPP levels and internalizing behavior in each time point; (B) association between BDCPP levels and externalizing behavior in each time point; (C) association between DPHP and internalizing behavior in each time point; (D) association between DPHP levels and externalizing behavior in each time point. Effect estimates and 95%-confidence intervals are presented per an increase of one SD in the metabolite levels. All associations in adjusted models 1 were adjusted for maternal BMI and age at enrollment, child’s ancestry, and SES sum score. All associations in “Adjusted model 1 + creatinine” were additionally adjusted for creatinine concentration. The sample sizes for the study groups with outcomes measured at 24, 42, and 60 months were 459, 662, and 678, respectively. The sample sizes for models with creatinine concentration were 457, 659, and 675, respectively.

Specifically, a one SD increase in the natural logs of BDCPP concentrations were associated with 1.26-point higher internalizing t-scores (95% CI: 0.02, 2.50) and 1.40-point higher externalizing t-scores (95% CI: 0.34, 2.46). Similarly, a SD increase in the natural log of DPHP concentrations was associated with 1.00-point higher internalizing behaviors scores (95% CI: -0.12, 2.14) and 1.20-point higher externalizing behaviors scores (95% CI: 0.23, 2.16) after adjusting for maternal BMI, maternal age, ancestry, and SES (Supplemental Table 2, Figure 1).

When urinary creatinine concentration was included in the adjusted models, most of the effect estimates were slightly attenuated, and association at 24 months were no longer statistically significant for CBCL (Figure 1,Supplemental Table 2).

At later timepoints (42 and 60 months), most associations between prenatal OPFR metabolite levels and internalizing and externalizing behavior were mostly null (Supplemental Table 2, Figure 1).

Only DPHP concentrations remained positively associated with internalizing behavior at 60 months (Figure 1C). A SD increase in the natural logs of DPHP concentrations were associated with 0.97-point higher internalizing behaviors scores (95% CI: -0.03, 1.96) in adjusted models and 1.10-points higher scores (95% CI: 0.01, 2.18) after additionally adjusting for creatinine.

### Sensitivity analyses

The association results from the full analysis sample were similar to the associations observed in the subset of individuals with urinary OPFR metabolite measurements during the second trimester pregnancy at three time points (Figure 2). Effect sizes and directions were consistent across all models.

**Figure 2.**
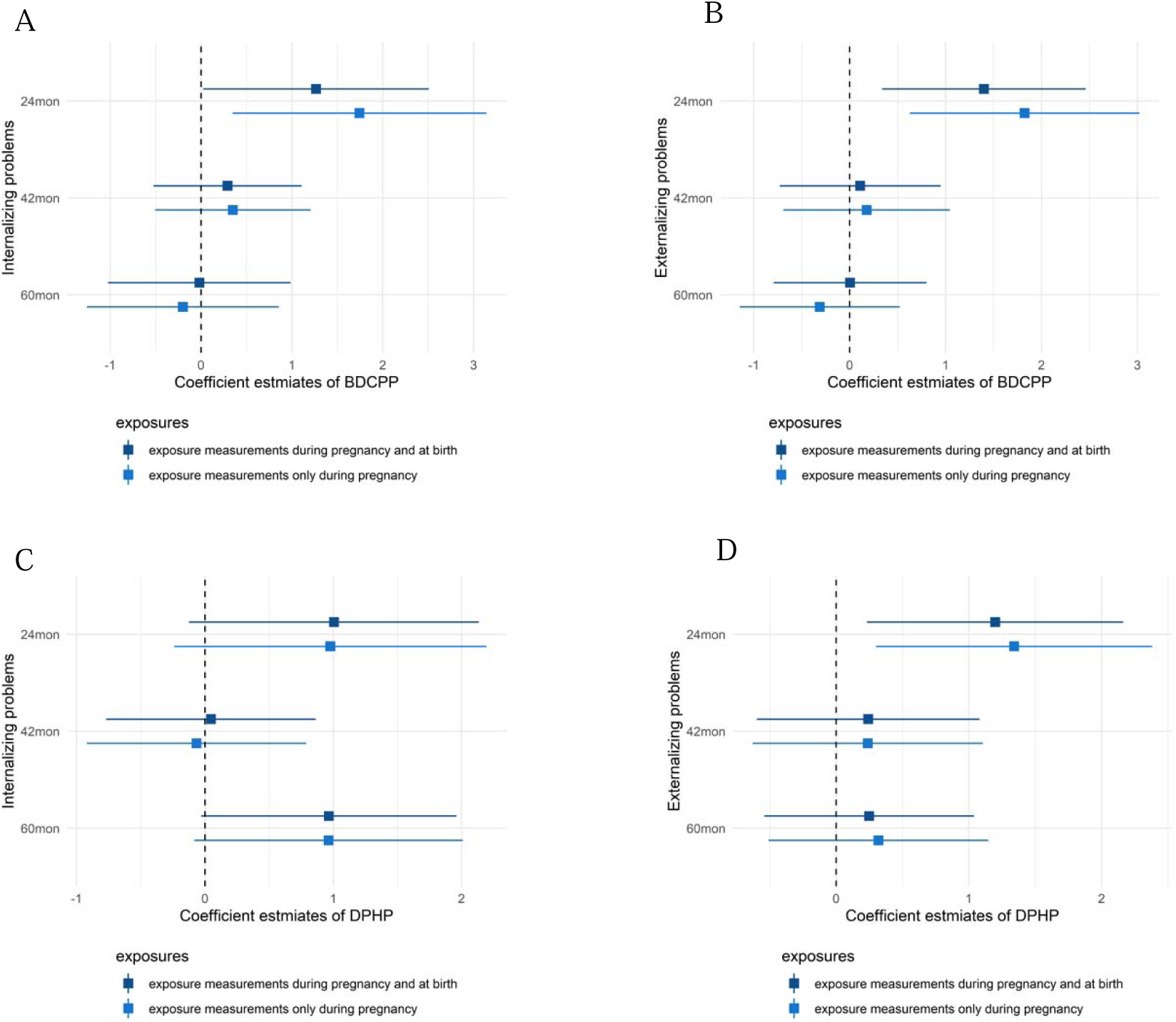
Association between prenatal OPFR metabolite levels (BDCPP and DPHP) and internalizing and externalizing behaviors at 24, 42, and 60 months among full study sample and individuals with exposure measurements during pregnancy in Drakenstein Child Health Study from 2012 to 2015, Cape Town, South Africa. (A) Association between BDCPP levels and internalizing behavior in each time point; (B) association between BDCPP levels and externalizing behavior in each time point; (C) association between DPHP levels and internalizing behavior in each time point; (D) association between DPHP levels and externalizing behavior in each time point. Effect estimates and 95%-confidence intervals are presented per an increase of one SD in the metabolite levels. All associations were adjusted for maternal BMI and age at enrollment, child’s ancestry, and SES sum score. Sample sizes for subsamples at 24, 42, and 60 months were 413, 604, and 619, respectively.

Since the detection frequency of OPFR metabolites in urine samples were relatively low, we dichotomized their levels into detected versus not detected. Models for BDCPP detection and neurobehavioral outcomes showed similar effect sizes and same directions with main results. While models for DPHP detection and internalizing/externalizing behaviors were not significant, the directions in models were consistent with our main results (Figure 1 and Figure 3).

**Figure 3.**
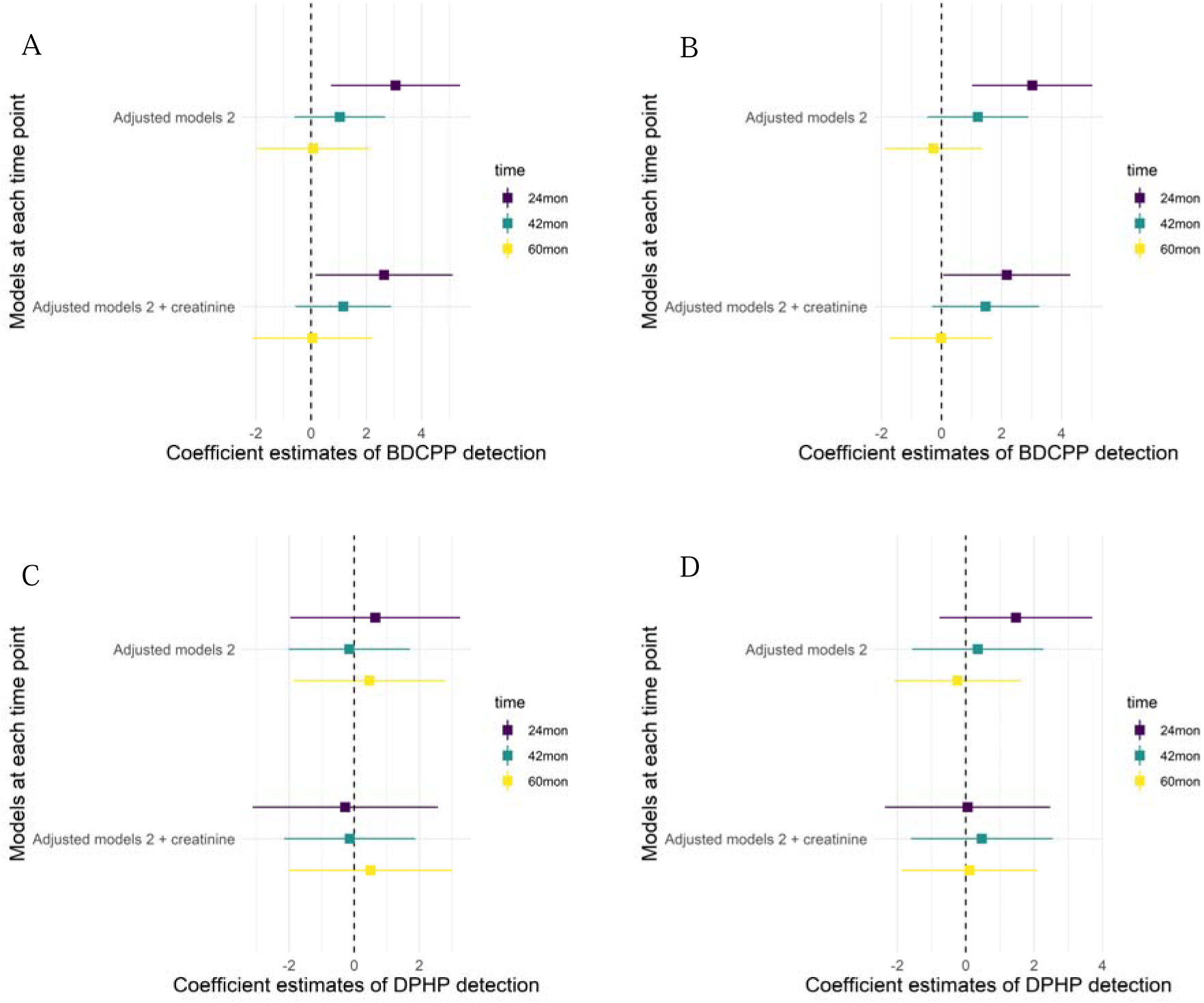
Association between prenatal OPFR metabolites detection and internalizing and externalizing behaviors at 24, 42, and 60 months in Drakenstein Child Health Study from 2012 to 2015, Cape Town, South Africa. (A) Association between BDCPP detection and internalizing behavior in each time point; (B) association between BDCPP detection and externalizing behavior in each time point; (C) Association between DPHP detection and internalizing behavior in each time point; (D) association between DPHP detection and externalizing behavior in each time point. Effect estimates and 95%-confidence intervals are presented per an increase of one unit in the metabolite detection. All associations in adjusted models 2 were adjusted for maternal BMI and age at enrollment, child’s ancestry, and SES sum score. All associations in “Adjusted model 2 + creatinine” were additionally adjusted for creatinine concentration. The sample sizes for the study groups with outcomes measured at 24, 42, and 60 months were 459, 662, and 678, respectively. The sample sizes for models with creatinine concentration were 457, 659, and 675, respectively.

Effect modification analysis by child sex showed that associations between OPFR metabolite levels and externalizing behavior were stronger among males than females across all timepoints and exposures, but the effect modification was only significant for DPHP and externalizing behavior at 42 months (Figure 4). Associations between OPFRs urinary metabolite levels and internalizing behavior did not differ by child sex.

**Figure 4.**
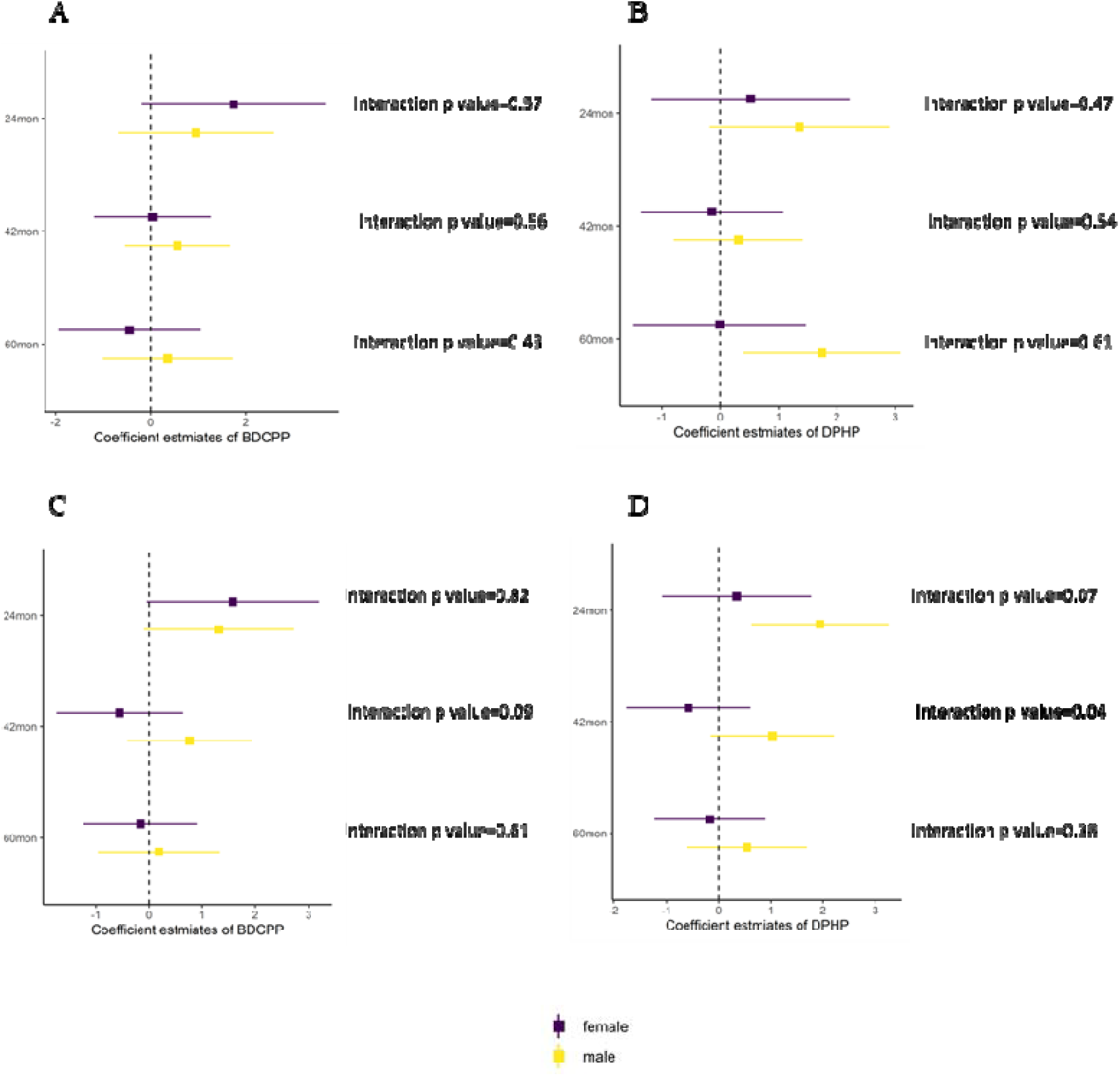
Association between prenatal OPFR metabolite levels (BDCPP and DPHP) and internalizing and externalizing behaviors at 24, 42, and 60 months among full study sample, stratified by infant sex in Drakenstein Child Health Study from 2012 to 2015, Cape Town, South Africa. (A) Association between BDCPP levels and internalizing behavior in each time point; (B) association between BDCPP levels and externalizing behavior in each time point; (C) association between DPHP levels and internalizing behavior in each time point; (D) association between DPHP and externalizing behavior in each time point. Effect estimates and 95%-confidence intervals are presented per an increase of one SD in the metabolite levels. All associations were adjusted for maternal BMI and age at enrollment, child’s ancestry, and SES sum score.

## Discussion

In this prospective cohort study of 705 mother-child pairs from South Africa, higher maternal OPFR urinary metabolite levels pregnancy and at birth were associated with higher scores of behaviors in children at the age of two years. However, the association was attenuated in later childhood, with the exception of the association between DPHP urinary levels and internalizing behaviors at 60 months. We observed significant associations between prenatal BDCPP levels and children’s internalizing and externalizing behavior at two years old. Furthermore, prenatal urinary DPHP levels were significantly associated with externalizing behaviors in children at two years of age and with internalizing behaviors at 5 yrs. These findings suggest that prenatal exposure to specific OPFRs may influence the early development of behavioral problems which could impact the children’s long-term emotional and psychological well-being. To our knowledge, this study was the first investigation of the transgenerational neurotoxic effect of OPFRs (assessed using maternal urinary metabolites) among children in LMICs.

When compared to OPFR metabolite levels reported in previous studies of pregnant women, the concentrations of a BDCPP in our study were similar to levels found in women from China, ^14,28^ Canada, ^29^ and Norway, ^30^ but lower than those reported in some regions of the United States. ^15,31^ The concentrations of DPHP in our study were similar to that of found in women from the United States ^15,31^ but slightly higher than levels observed in women from Norway ^30^ and China. ^14^ These comparisons demonstrate the ubiquitous and global exposure to OPFRs among pregnant women. However, the detection frequencies observed in our study were generally lower than those reported in the aforementioned studies. With the exception of investigations conducted in Norway (52%) ^30^ and Canada (29%) ^29^, studies from the United States ^15,16,31,32^ and China ^14,33^ consistently reported BDCPP detection frequencies exceeding 70%, with some exceeding 90%. Similarly, all prior studies documented high detection frequencies for DPHP (>70%). Although our study identified DPHP in 71% of samples, most previous investigations reported substantially higher frequencies, in some cases surpassing 95%. ^14,29,30,32,33^

To our knowledge, eight other studies have explored the associations between OPFR metabolite levels and childhood neurodevelopment and behaviors. ^14–18,28,33,34^ Six of these were longitudinal studies investigating the relationship between maternal OPFR metabolite levels during pregnancy and neurodevelopmental outcomes in children from 1 to 7 years old, ^14–16,28,33,34^ while two cross-sectional studies measured OPFR metabolites in child urine and investigated their association with social behaviors. ^17,18^

Most studies have reported associations between BDCPP levels and neurodevelopment, consistent with our findings. ^14,16,32,33^ Comparable results were observed in Wuhan (N = 184) ^14^ and Shandong, China (N = 270) ^33^, where BDCPP levels correlated with lower cognitive scores and neurodevelopmental delays measured in offspring aged 1 and 2 years. However, findings in the United States were different. Doherty et al. reported no association between prenatal BDCPP levels and early learning or communicative development at 36 months (N = 149). ^16^ Similarly, the MADRES cohort (N = 329) ^32^ found no evidence linking prenatal BDCPP levels with gross or fine motor delays in offspring aged 6 to 18 months.

In contrast, the association between prenatal DPHP levels and childhood neurodevelopmental deficits was more consistent across studies, aligning with our results. ^15,28,32^ Although Doherty et al. ^16^ did not identify a significant relationship, the Ma’anshan (N = 1781) ^28^ and CHAMACOS birth cohort studies (N = 310) ^15^ revealed significant negative associations of DPHP exposure during pregnancy on working memory in children aged 3 to 7 years, reinforcing our findings on prenatal DPHP concentration and early childhood neurobehavioral impairments. The Ma’anshan birth cohort study also demonstrated a negative association of first trimester DPHP levels exposure on full-scale intelligence quotient in children aged 3 to 6 years. While there was no evidence supporting an association between prenatal BDCPP levels and developmental delays, the MADRES cohort found a positive association between prenatal DPHP levels and fine motor delays among offspring aged 6 to 18 months. ^32^

Our study identified a stronger association between prenatal DPHP levels and externalizing behaviors in males compared to females. Child’s sex significantly modified the effect of prenatal DPHP levels on externalizing behaviors at 42 months. A potential explanation for this finding is that externalizing behaviors are more common in boys than in girls. In line with previous studies, ^35,36^ male children in the DCHS had higher average scores and variation of t scores in externalizing problems (t-scores at 42 months = 42.9, SD = 10.9) compared to females (t-scores at 42 months = 40.8, SD = 10.5). It is well known that chemicals exposures, including OPFRs, can affect child sex-specific neurodevelopmental and behavioral outcomes differently. ^14,32,37^ For example, the MADRES cohort found a significant sex interaction with BDCPP levels and internalizing and total problems measured by CBCL ^34^ as well as with adverse fine motor movement assessed with the Ages and Stages Questionnaire-3 (ASQ-3). ^32^ Another study from Wuhan, China, provided evidence of sex-specific patterns in the relationship between BDCPP levels measured during the first trimester and Psychomotor Developmental Index (PDI) and Mental Developmental Index (MDI) scores. ^14^ The low detection frequency of BDCPP in our urine samples might explain why we did not find the same significant sex differences in the association between prenatal BDCPP levels and neurobehavioral problems. These findings highlight the need for subsequent studies focused on the sex-modification effects related to OPFR exposures.

This study has several strengths. Firstly, it was based on a birth cohort study with a prospective design and repeated measurements of child behavior, enabling the evaluation of behavioral outcomes from age 2 to 5 years. Secondly, it is the first study to investigate these associations within an African population in the context of LMICs, providing valuable insights into this underrepresented group. Thirdly, we included various potential confounding variables in our analyses to reduce the risk of confounding bias, including BMI and creatinine, which are both indicators of individual metabolism ^38,39^. Only two previous studies adjusted for creatinine concentrations using the chemical-creatinine ratio. ^28,33^ However, this study also has several limitations. Firstly, we did not have repeated measures of exposure throughout the entire pregnancy. Given the short biological half-lives of OPFRs, a single spot urine sample may not fully reflect the overall prenatal exposure to OPFRs. However, the consistency of our results between measurements taken during pregnancy versus at birth, suggests that this limitation might not diminish the validity of our results. Nevertheless, the sensitivity analyses demonstrated that models using dichotomized OPFR metabolite levels yielded results consistent with those using continuous measures. Therefore, the limited detection frequency is unlikely to substantially affect the validity of the results. Thirdly, we did not include data of biological samples from the children themselves, meaning we were unable to assess postnatal exposure, which may also influence child neurodevelopmental outcomes. Finally, our analysis focused solely on the individual effects of OPFR metabolites, without accounting for potential interactions or combined effects of exposure to multiple OPFRs.

Overall, we observed a positive association between prenatal OPFR metabolite levels and neurobehavioral problems among children at age two in South Africa, raising concerns about the impact of prenatal chemical exposures on early brain development. This also underscores the need for increased awareness and the development of interventions aimed at promoting healthier developmental outcomes for children. Given the widespread use and environmental presence of OPFRs, further research is essential to better understand the underlying mechanisms of neurotoxicity and the long-term effects of OPFR exposures on child development.

## Data Availability

All data produced in the present study are available upon reasonable request to the authors

## Description of conflicts of interest

Dan J Stein has received consultancy honoraria from Discovery Vitality, Kanna, L’Oreal, Lundbeck, Orion, Servier, Seaport Therapeutics, Takeda, Vistagen, and Wellcome.

## Data sharing

The DCHS is committed to the principle of data sharing. De-identified data will be made available to requesting researchers as appropriate. Requests for collaborations to undertake data analysis are welcome. More information can be found on our website [http://www.paediatrics.uct.ac.za/scah/dclhs].

## Acknowledgements

We thank the staff of the DCHS, the staff of the Western Cape Health Dept for their support of the study and the families and children in the DCHS.

The Mount Sinai HHEAR Targeted Analysis Laboratory expresses appreciation to Ravikumar Jagani, Jasmin Chovatiya, and Shirisha Yelamanchili for conducting lab assays on urine samples for organophosphorus flame-retardant metabolites and creatinine.

---

**Supplemental Figure 1.**
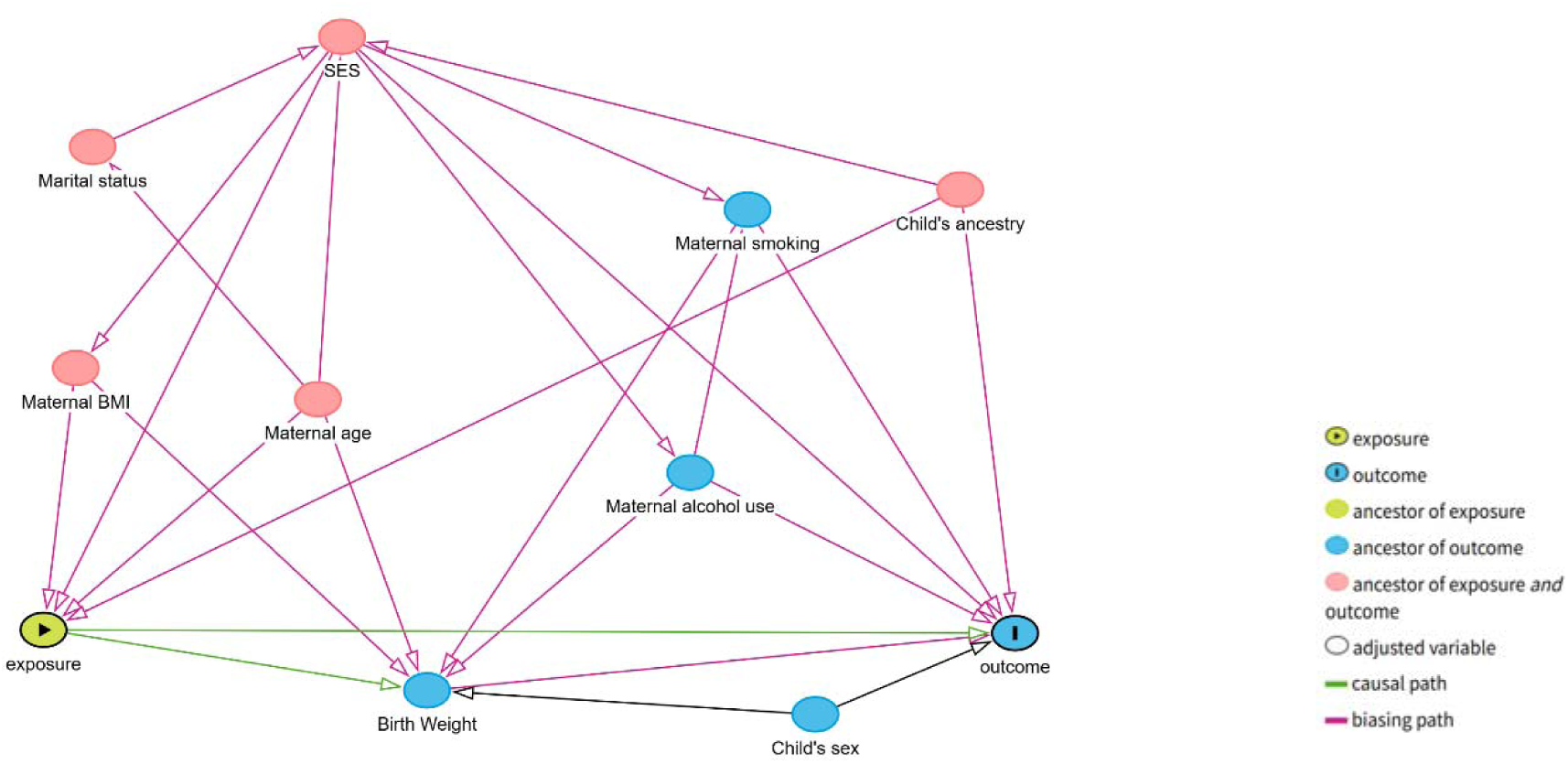
Directed acyclic graphs (DAG) of the association between prenatal OPFRs exposure and childhood behaviors in Drakenstein Child Health Study from 2012 to 2015, Cape Town, South Africa *Exposure: Prenatal OPFRs exposure; outcome: Childhood behaviors*.

**Supplemental Table 1.** Concentrations of OPFRs metabolites levels measured during pregnancy and at birth (ng/ml) with CBCL measurements at 24, 42 and 60 months in Drakenstein Child Health Study from 2012 to 2015, Cape Town, South Africa.

| OPFRs | 24 months |  | 42 months |  | 60 months |  |
| --- | --- | --- | --- | --- | --- | --- |
|  | Pregnancy<br>(N=413) | Birth<br>(N=46) | Pregnancy<br>(N=604) | Birth<br>(N=58) | Pregnancy<br>(N=619) | Birth<br>(N=59) |
| BDCPP,<br>median<br>(IQR) | 0.24 (0.71) | 0.73<br>(2.43) | 0.29 (0.95) | 0.70<br>(2.25) | 0.30 (0.99) | 0.74<br>(2.40) |
| DPHP,<br>median<br>(IQR) | 1.00 (1.7) | 1.60<br>(2.80) | 1.01 (1.56) | 1.60<br>(2.59) | 1.06 (1.65) | 1.63<br>(2.69) |

*Abbreviations: BDCPP: bis(1,3-dichloro-2-propyl) phosphate; DPHP: diphenyl phosphate.*

**Supplemental Table 2.** Unadjusted and adjusted linear regression models of the association between BDCPP and DPHP metabolites levels and childhood behaviors in Drakenstein Child Health Study from 2012 to 2015, Cape Town, South Africa.

|  | Unadjusted models |  | Adjusted models 1 |  | Adjusted models 2 |  |
| --- | --- | --- | --- | --- | --- | --- |
|  | Standardized BDCPP | Standardized DPHP | Standardized BDCPP | Standardized DPHP | Standardized BDCPP | Standardized DPHP |
| **24 m** |  |  |  |  |  |  |
| In. | 1.16<br>(-0.06, 2.39) | 0.89<br>(-0.21, 2.00) | 1.26*<br>(0.02, 2.50) | 1.00<br>(-0.12, 2.14) | 1.06<br>(-0.21, 2.34) | 0.70<br>(-0.57, 1.96) |
| Ex. | 1.37*<br>(0.32, 2.42) | 1.07*<br>(0.12, 2.02) | 1.40*<br>(0.34, 2.46) | 1.20*<br>(0.23, 2.16) | 1.06<br>(-0.02, 2.14) | 0.61<br>(-0.47, 1.68) |
| **42 m** |  |  |  |  |  |  |
| In. | 0.13<br>(-0.67, 0.94) | 0.08<br>(-0.73, 0.88) | 0.29<br>(-0.52, 1.11) | 0.05<br>(-0.77, 0.86) | 0.31<br>(-0.53, 1.15) | 0.06<br>(-0.83, 0.95) |
| Ex. | -0.06<br>(-0.90, 0.77) | 0.30<br>(-0.54, 1.13) | 0.11<br>(-0.73, 0.95) | 0.24<br>(-0.60, 1.08) | 0.15<br>(-0.71, 1.02) | 0.34<br>(-0.57, 1.26) |
| **60 m** |  |  |  |  |  |  |
| In. | -0.02<br>(-1.02, 0.97) | 1.10*<br>(0.11, 2.08) | -0.02<br>(-1.02, 0.99) | 0.97<br>(-0.03, 1.96) | -0.03<br>(-1.07, 1.01) | 1.10*<br>(0.01, 2.18) |
| Ex. | -0.09<br>(-0.89, 0.70) | 0.32<br>(-0.47, 1.12) | 0.00<br>(-0.79, 0.80) | 0.25<br>(-0.54, 1.04) | 0.11<br>(-0.71, 0.94) | 0.45<br>(-0.41, 1.31) |

\* $p < 0.05$

